# Assessment of Sociodemographic Factors Associated with Time to Self-reported COVID-19 Infection Among a Large Multi-Center Prospective Cohort Population in the Southeastern United States

**DOI:** 10.1101/2023.10.20.23297306

**Authors:** Andrew J. Beron, Joshua O. Yukich, Andrea A. Berry, Adolfo Correa, Joseph Keating, Matthew Bott, Thomas F Wierzba, William S. Weintraub, DeAnna J. Friedman-Klabanoff, Morgana Mongraw-Chaffin, Michael A. Gibbs, Yhenneko J. Taylor, Patricia J. Kissinger, Devin Hayes, John S. Schieffelin, Brian K. Burke, Richard A. Oberhelman

## Abstract

**Objective:** We aimed to investigate sociodemographic factors associated with self-reported COVID-19 infection.

**Methods:** The study population is a multicenter prospective cohort of adult volunteers recruited from healthcare systems located in the mid-Atlantic and southern United States. Between April 2020 and October 2021 participants completed daily online questionnaires about symptoms, exposures, and risk behaviors related to COVID-19, including self-reports of positive SARS CoV-2 detection tests and COVID-19 vaccination. Analysis of time from study enrollment to self-reported COVID-19 infection used a time-varying mixed effects Cox-proportional hazards framework.

**Results:** Overall, 1,603 of 27,214 study participants (5.9%) reported a positive COVID-19 test during the study period. The adjusted hazard ratio demonstrated lower risk for women, those with a graduate level degree, and smokers. A higher risk was observed for healthcare workers, those aged 18-34, those in rural areas, those from households where a member attends school or interacts with the public, and those who visited a health provider in the last year.

**Conclusions:** Increased risk of self-reported COVID-19 was associated with specific demographic characteristics, which may help to inform targeted interventions for future pandemics.

## INTRODUCTION

The COVID-19 pandemic has created and widened existing health disparities within US society. Epidemiologic data collected during COVID-19 surveillance provides important insights on at-risk populations including differential access to information and infection control measures, in particular social distancing, masking, and new vaccines to guide future pandemic preparedness. Regional studies carried out by local U.S. healthcare and public health organizations, as well as national cross-sectional studies^1,2,3^ have found higher incidence in ethnic and race-based minority populations and certain age groups. ^4^ Nonetheless, there is a paucity of comprehensive risk factor analyses beyond demographic characteristics that examine other factors such as occupation, self-reported health conditions, health behaviors, and household characteristics.

The COVID-19 Community Research Partnership (CCRP) was a multi-state cohort study designed to monitor the evolution of the pandemic in a large population with both syndromic surveillance and periodic testing for serologic evidence of infection. The goal of the CCRP was to generate data to inform ongoing public health responses to COVID-19 as well as future pandemics by recruiting a diverse cohort of patients and community members. The methods and purpose have been described elsewhere. ^5^

This large, multicenter study provided an opportunity to further examine population-based risk factors for COVID-19 to identify characteristics of subgroups most likely to become infected. As such, the purpose of this paper is to investigate factors associated with time from enrollment in follow-up to a self-reported COVID-19 infection. Herein we report findings of our risk factor analysis comparing hazard rates in subgroups based on individual and household characteristics.

## MATERIALS AND METHODS

Participants were drawn from the prospective, multi-site, CCRP COVID-19 surveillance cohort study, a convenience sample of patients and healthcare workers in ten healthcare systems from the Mid-Atlantic and southeastern US. We recruited adults through patient portals or email from the following health systems and institutions: Wake Forest Baptist Health, Atrium Health, Wake Med, New Hanover Regional Medical Center, Vidant Health, Campbell University, Tulane University affiliated partner systems, University of Mississippi, University of Maryland Medical System, and Medstar Health. Adults aged 18 years and older were eligible to participate if they were a patient or employee of a participating healthcare system. Only participants who reported residing in Maryland, Virginia, Washington D.C., North Carolina, South Carolina, Mississippi, and Louisiana were included. Participants who reported a prior positive test for COVID-19 on enrollment were excluded.

Study procedures included daily online questionnaires for all participants, extracting electronic health records (EHR) for those who were patients at a participating health system, and periodic at-home serological testing for a subset of participants. The Wake Forest Baptist Health Institutional Review Board (IRB), which served as the central IRB for this study, approved the study protocol. Internet-based informed consent through secure patient portals was obtained from study participants prior to any study procedures. The study is registered with ClinicalTrials.gov, NCT04342884. Enrollment began April 8, 2020, and ended on April 29, 2022.

### Participant data

Participants in the study completed daily online questionnaires about symptoms, exposures, and risk avoidance behaviors related to COVID-19, including self-reports of recent positive SARS CoV-2 detection tests (herein described as “self-reported COVID-19”). Respondents also reported history of COVID-19 vaccination (date of receipt, product, dose 1 or dose 2, participation in a clinical trial). Race was defined based on responses to the initial study enrollment questionnaire, with options listed as 1) Black or African American, 2) Asian, 3) Hispanic or Latino, 4) White (not Hispanic/Latino), 5) American Indian or Alaskan Native, and 6) Mixed Ethnicity. Participants were invited to complete two online supplemental questionnaires, one that was focused on the individual and another that was focused on the individual’s household, to provide more detailed information on demographic characteristics, occupations, self-reported health conditions, health behaviors, and household characteristics. Supplemental questionnaires were sent to all actively enrolled participants in May 2021, and subsequent newly enrolled study participants received the surveys within a month of starting the study.

The study sought to examine correlations between the occupational characteristics of participants and the primary outcome. The National Institute for Occupational Safety and Health (NIOSH), a United States federal agency responsible for conducting research and making recommendations for the prevention of work-related injury and illness, has characterized workplace exposure to SARS-CoV-2 in hundreds of non-health care occupations using metrics from O*NET, a national database with information on occupational characteristics, together with input from experts in occupational safety and health. Based on these data, occupations are categorized in three risk levels (i.e., high, medium, low) using the SARS-CoV-2 Occupational Exposure Matrix (SOEM) system. ^6^ SOEM exposure categories were based on three factors identified as contributing to increased risk of exposure in the workplace: whether an occupation involves routine in-person interaction with the public (*Public Facing*), working indoors (*Working Indoors*), and working in close physical proximity to others, either co-workers or the public (*Close Proximity*). Since health care workers were over-represented in the study population, the high exposure group was divided into two groups for data analysis, i.e., high-healthcare, and high-non healthcare.

### Data analysis

Data from participants who completed both supplemental questionnaires were analyzed to determine demographic, occupational, health-related, and behavioral correlates of self-reported SARS CoV-2 infection. Descriptive statistics were produced by cross tabulation and covariates for model inclusion were first checked for collinearity using pairwise correlation. Analysis followed a time-varying mixed effects Cox-proportional hazards framework, with a shared frailty at the level of recruitment site to account for homogeneity within each site/health network and to account for intraclass correlation in the outcome of interest (self-reported COVID-19 infection) within health networks.

Hazard Ratios and 95% confidence intervals are reported as unadjusted estimates and as adjusted for all covariates included in the final model (Table 2). Survival-Curves are also presented using the Kaplan-Meier Product Limit estimator for selected covariates.

Two time-varying covariates were included in the categorical hazard analysis: 1) county level 7-day average COVID-19 incidence data updated daily and published by the New York Times in 2020 and 2021 ^7^ and 2) COVID-19 vaccination status of participants. Participants were considered vaccinated after they reported receiving their first vaccination dose of any COVID-19 vaccine.

## RESULTS

A total of 69,714 participants were enrolled in the CCRP study. Of those, 42,701 (61.3%) completed the individual adult supplemental survey and 31,642 (45.4%) completed the household supplemental survey. Just under thirty thousand (29,973 (43.0%)) participants completed both the individual adult and household supplemental surveys. Participants that reported residing outside of Maryland, Virginia, Washington D.C., North Carolina, South Carolina, Mississippi, and Louisiana, as well as participants that were participating in clinical trials were excluded, leaving a total of 27,214 participants (39% of the total number of CCRP study participants) in the analytic population.

Overall, 1,603 of a total of the 27,214 study participants (5.9%) reported that they had a positive test for COVID-19 between enrollment and the end of October 2021 (Table 1). The study population was predominantly female (71.4%) and White/non-Hispanic (88.3%; Table 1). Most participants lived in counties classified as urban (56.1%) and 96.9% had at least some college education with a large proportion holding graduate level degrees (47.9%). The study population was affluent (54.1% had a household income over $100,000). Data was not available for SOEM category designation for 32% of subjects, but among the other participants there was an even distribution between the four SOEM exposure groups (low, medium, high-non healthcare, and high-healthcare). The networks with the largest numbers of participants included Wake Forest (32.6%), MedStar (26.7%), and Atrium (17.1%).

**Table 1.** Characteristics at baseline of participants enrolled in the COVID-19 Community Research Panel included for analysis by Self-Reported COVID-19 Status.

| Variable | Overall number of participants* | No* | Yes* | Percentage with Self-Reported COVID-19 Diagnosis |
| --- | --- | --- | --- | --- |
| Total subjects | 27,214 | 25,611 | 1,603 | 5.9 |
| Female | 19,443 (71.4) | 18,266 (71.3) | 1,177 (73.4) | 6.1 |
| Age group |  |  |  |  |
| 18-34 | 2,911 (10.7) | 2,675 (10.4) | 236 (14.7) | 8.1 |
| 35-49 | 7,395 (27.2) | 6,808 (26.6) | 587 (36.6) | 7.9 |
| 50-64 | 9,388 (34.5) | 8,832 (34.5) | 556 (34.7) | 5.9 |
| 65+ | 7,520 (27.6) | 7,296 (28.5) | 224 (14.0) | 3 |
| Race/Ethnicity |  |  |  |  |
| White NH | 24,032 (88.3) | 22,582 (88.2) | 1,450 (90.5) | 6 |
| Black NH | 1,439 (5.3) | 1,377 (5.4) | 62 (3.9) | 4.3 |
| Hispanic | 604 (2.2) | 563 (2.2) | 41 (2.6) | 6.8 |
| Asian | 488 (1.8) | 470 (1.8) | 18 (1.1) | 3.7 |
| Other | 651 (2.4) | 619 (2.4) | 32 (2.0) | 4.9 |
| County Classification |  |  |  |  |
| Rural | 5,587 (20.5) | 5,184 (20.2) | 403 (25.1) | 7.2 |
| Suburban | 6,355 (23.4) | 5,950 (23.2) | 405 (25.3) | 6.4 |
| Urban | 15,271 (56.1) | 14,476 (56.5) | 795 (49.6) | 5.2 |
| Health Care Worker lives in Household | 6,006 (22.1) | 5,441 (21.2) | 565 (35.2) | 9.4 |
| Education Level of Respondent |  |  |  |  |
| Hs or less | 845 (3.1) | 782 (3.1) | 63 (3.9) | 7.5 |
| Some college | 2,756 (10.1) | 2,564 (10.0) | 192 (12.0) | 7 |
| Associate | 2,090 (7.7) | 1,882 (7.3) | 208 (13.0) | 10 |
| Bachelor | 8,483 (31.2) | 7,927 (31.0) | 556 (34.7) | 6.6 |
| Graduate | 13,040 (47.9) | 12,456 (48.6) | 584 (36.4) | 4.5 |
| History of Influenza Vaccination | 26,338 (96.8) | 24,800 (96.8) | 1,538 (95.9) | 5.8 |
| Time Since Last Primary Care Visit |  |  |  |  |
| <1 Year | 22,590 (83.0) | 21,232 (82.9) | 1,358 (84.7) | 6 |
| 1-2 Years Ago | 3,311 (12.2) | 3,137 (12.2) | 174 (10.9) | 5.3 |
| 2-5 Years Ago | 912 (3.4) | 871 (3.4) | 41 (2.6) | 4.5 |
| >5 Years Ago | 401 (1.5) | 371 (1.4) | 30 (1.9) | 7.5 |
| Current Tobacco Use of Respondent | 1,485 (5.5) | 1,410 (5.5) | 75 (4.7) | 5.1 |
| Number of Concurrent Medical Conditions Reported |  |  |  |  |
| None | 9,073 (33.3) | 8,520 (33.3) | 553 (34.5) | 6.1 |
| One | 8,961 (32.9) | 8,431 (32.9) | 530 (33.1) | 5.9 |
| Two | 5,603 (20.6) | 5,294 (20.7) | 309 (19.3) | 5.5 |
| 3 or More | 3,577 (13.1) | 3,366 (13.1) | 211 (13.2) | 5.9 |
| Household Annual Income |  |  |  |  |
| <50k | 2,749 (10.1) | 2,595 (10.1) | 154 (9.6) | 5.6 |
| 50-100k | 7,061 (25.9) | 6,559 (25.6) | 502 (31.3) | 7.1 |
| >100k | 14,732 (54.1) | 13,924 (54.4) | 808 (50.4) | 5.5 |
| No answer | 2,672 (9.8) | 2,533 (9.9) | 139 (8.7) | 5.2 |
| Someone in the Household Attends Class in Person | 6,911 (25.4) | 6,315 (24.7) | 596 (37.2) | 8.6 |
| Social Contact Worker in the Household | 10,279 (37.8) | 9,471 (37.0) | 808 (50.4) | 7.9 |
| Study Site |  |  |  |  |
| Atrium | 4,662 (17.1) | 4,221 (16.5) | 441 (27.5) | 9.5 |
| Campbell | 247 (0.9) | 238 (0.9) | 9 (0.6) | 3.6 |
| Maryland | 3,366 (12.4) | 3,298 (12.9) | 68 (4.2) | 2 |
| MedStar | 7,265 (26.7) | 7,100 (27.7) | 165 (10.3) | 2.3 |
| Mississippi | 182 (0.7) | 172 (0.7) | 10 (0.6) | 5.5 |
| New Hanover | 438 (1.6) | 408 (1.6) | 30 (1.9) | 6.8 |
| Tulane | 79 (0.3) | 71 (0.3) | 8 (0.5) | 10.1 |

| Variable | Overall number of participants* | No* | Yes* | Percentage with Self-Reported COVID-19 Diagnosis |
| --- | --- | --- | --- | --- |
| Vidant | 619 (2.3) | 585 (2.3) | 34 (2.1) | 5.5 |
| Wake Forest | 8,883 (32.6) | 8,132 (31.8) | 751 (46.8) | 8.5 |
| Wake Med | 1,473 (5.4) | 1,386 (5.4) | 87 (5.4) | 5.9 |
| SOEM Exposure Risk Categorization of Subjects Occupation |  |  |  |  |
| High - Healthcare | 5,080 (20.1) | 4,587 (19.4) | 493 (31.4) | 9.7 |
| High - Other | 5,179 (20.5) | 4,849 (20.5) | 330 (21.0) | 6.4 |
| Low | 4,316 (17.1) | 4,068 (17.2) | 248 (15.8) | 5.7 |
| Medium | 2,100 (8.3) | 1,948 (8.2) | 152 (9.7) | 7.2 |
| No Data | 8,577 (34.0) | 8,231 (34.8) | 346 (22.1) | 4 |

\* Values are all n (%) unless otherwise noted. No = Did not report COVID-19 infection during surveillance period; Yes = Did report COVID-19 infection during surveillance period

Adjusted and unadjusted hazard ratios for self-reported COVID-19 are shown in Table 2. Despite the fact that a higher proportion of women in the study population reported COVID-19 infections as compared to men (6.1% vs. 5.5%), the adjusted hazard ratio demonstrated lower risk for women (aHR= 0.87 [0.77-0.98]). When compared to the risk of COVID-19 among participants ages 18-34, significantly lower risk for infection were seen in all other age groups (vs. ages 35-49 aHR=0.80; vs. ages 50-64 aHR=0.63; vs. ages >65 aHR=0.45). No difference in risk of COVID-19 was seen based on race or ethnicity. A lower risk of self-reported infection for participants living in urban counties compared to rural counties seen in unadjusted analysis was attenuated and borderline significant in adjusted analysis (aHR=0.88 [0.77-1.00]). Educational level was strongly associated with risk, with significantly lower risk for participants with with a graduate level degree (aHR=0.57 [0.44-0.76]) compared to those with no college. As compared to participants who had seen their primary care provider within the past year, lower risk was observed for those whose last primary care visit was 1-2 years ago (aHR=0.84 [0.71-0.98]) or 2-5 years ago (aHR=0.68 [0.49-0.94]). Smokers had significantly lower risk of infection as compared to non-smokers (aHR=0.73 [0.58 - 0.92]). Risk did not correlate with the number of self-reported health conditions, but there was a borderline significant increase in risk for participants with 3 or more chronic health conditions as compared to those who reported no chronic conditions (aHR=1.18 [0.99 - 1.40]). Household income was not significantly associated with risk. Higher risk was observed in households where someone attended classes in-person (aHR=1.23 [1.11-1.37]) and in households where someone had occupational contact with the general public (aHR=1.24 [1.12 - 1.38]; Figure 1a). Receipt of at least one dose of COVID-19 vaccine was strongly protective (aHR=0.37; Figure 1b). In adjusted analyses based on SOEM occupational risk level, healthcare workers in the high-risk group had higher risk as compared to the high risk/non healthcare group (aHR=0.79 [0.68-0.92]), the medium risk group (aHR=0.82 [0.68-1.00]), and the low risk group (aHR=0.75 [0.64-0.89]; Figure 1c).

**Table 2.**
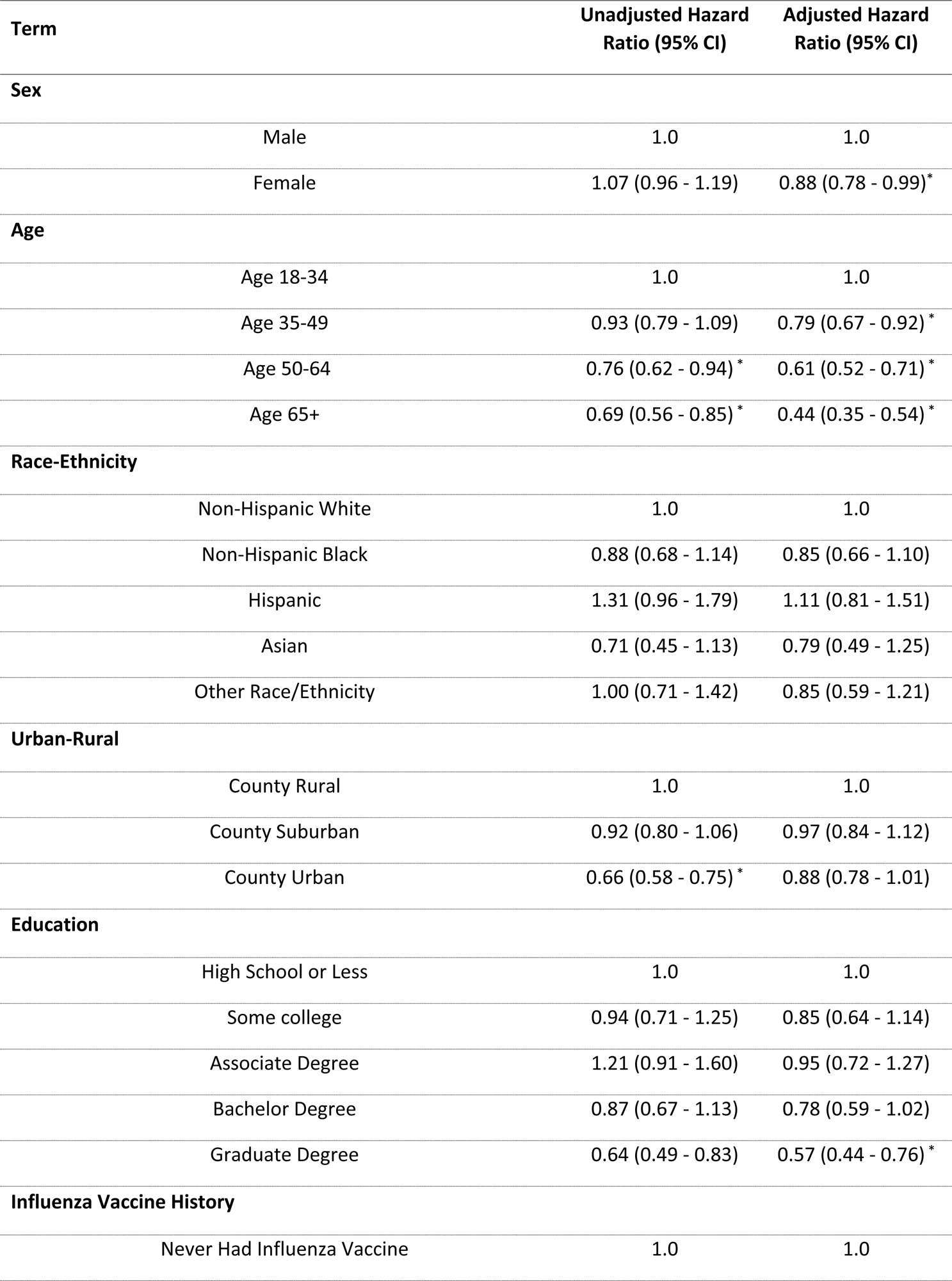
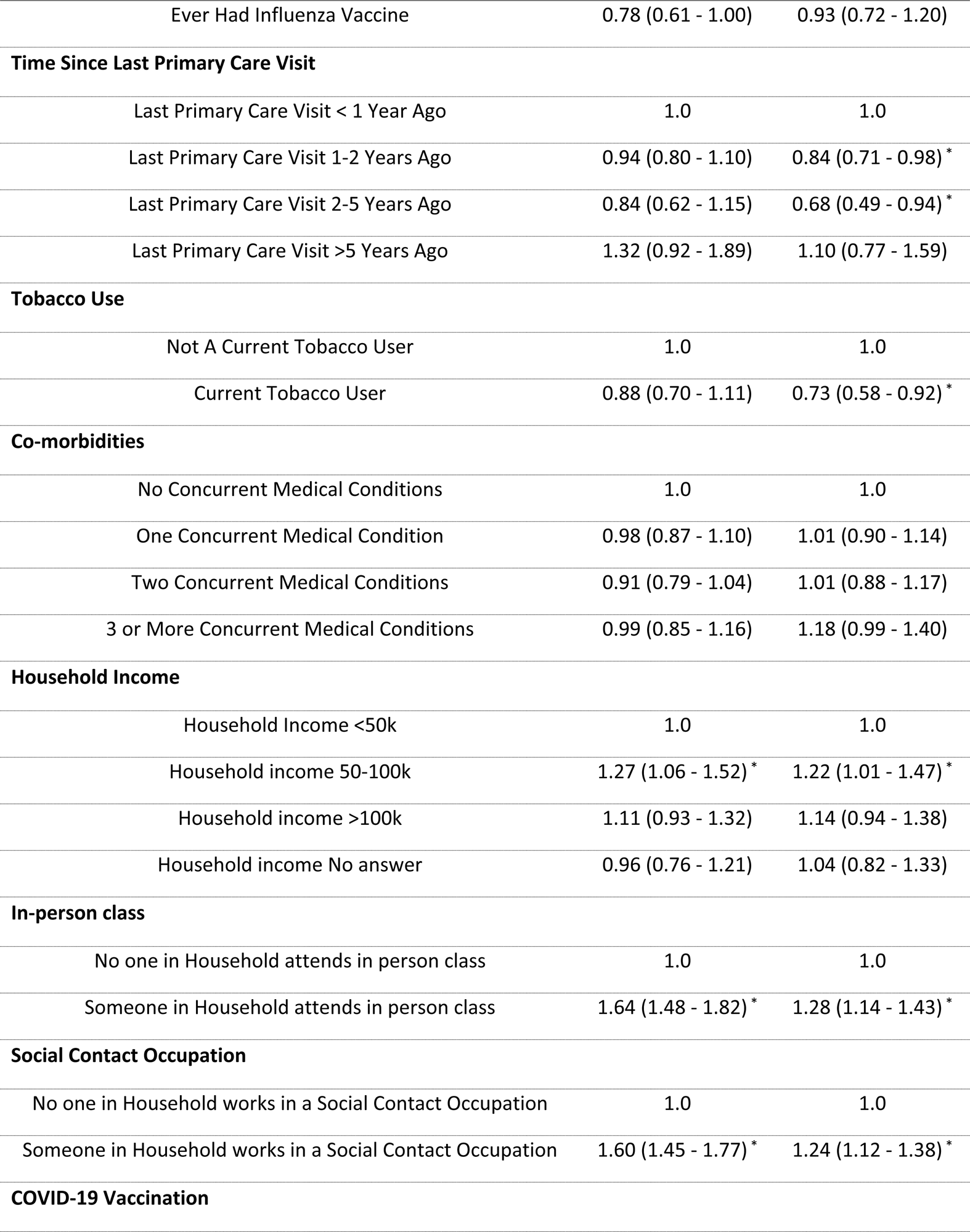
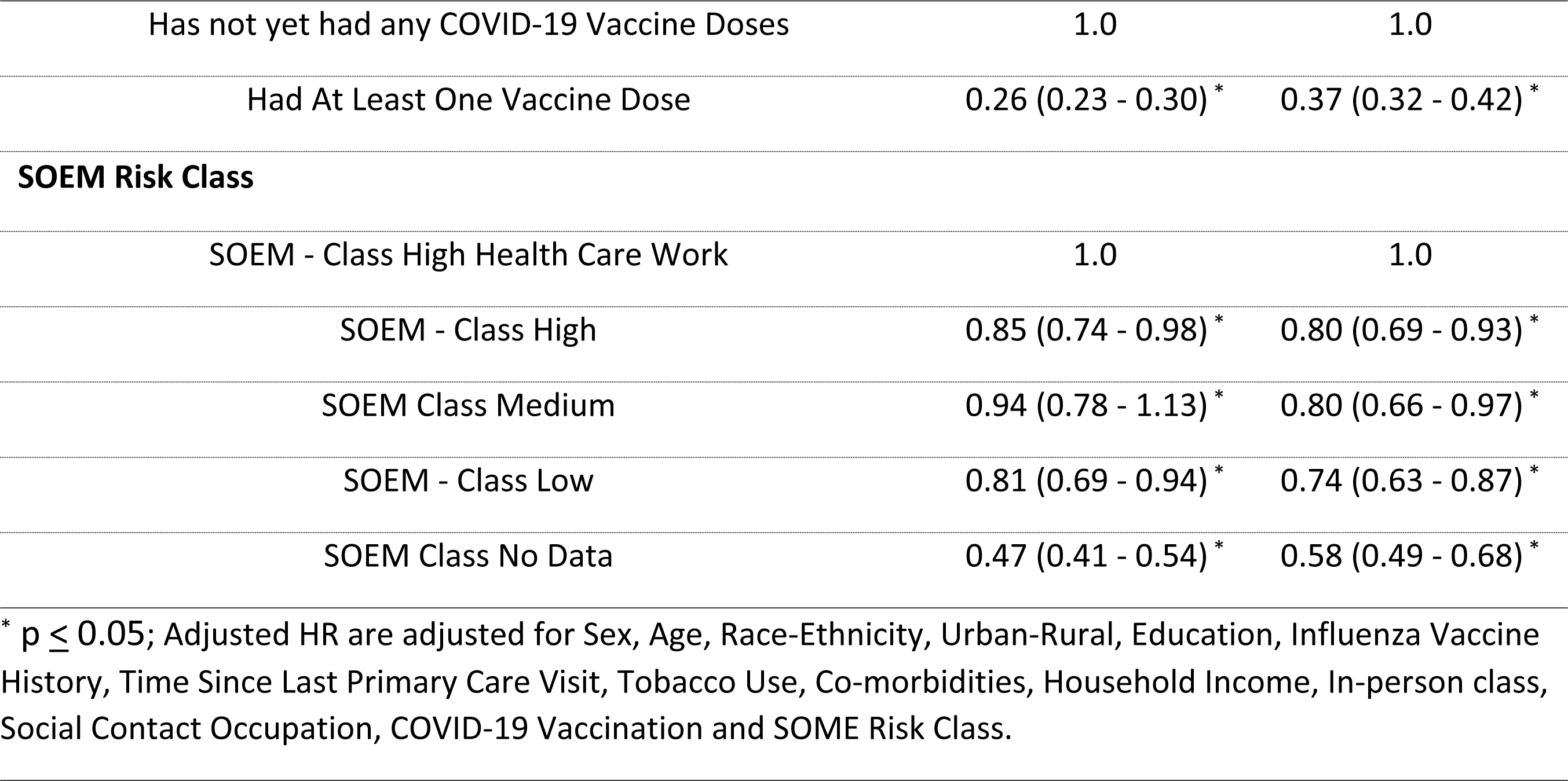
Determinants of Self-Reported COVID-19 Diagnosis following a time-varying mixed-effects Cox-proportional hazards model with shared frailty for health system.

| Term | Unadjusted Hazard Ratio (95% CI) | Adjusted Hazard Ratio (95% CI) |
| --- | --- | --- |
| **Sex** |  |  |
| Male | 1.0 | 1.0 |
| Female | 1.07 (0.96 - 1.19) | 0.88 (0.78 - 0.99)* |
| **Age** |  |  |
| Age 18-34 | 1.0 | 1.0 |
| Age 35-49 | 0.93 (0.79 - 1.09) | 0.79 (0.67 - 0.92)* |
| Age 50-64 | 0.76 (0.62 - 0.94)* | 0.61 (0.52 - 0.71)* |
| Age 65+ | 0.69 (0.56 - 0.85)* | 0.44 (0.35 - 0.54)* |
| **Race-Ethnicity** |  |  |
| Non-Hispanic White | 1.0 | 1.0 |
| Non-Hispanic Black | 0.88 (0.68 - 1.14) | 0.85 (0.66 - 1.10) |
| Hispanic | 1.31 (0.96 - 1.79) | 1.11 (0.81 - 1.51) |
| Asian | 0.71 (0.45 - 1.13) | 0.79 (0.49 - 1.25) |
| Other Race/Ethnicity | 1.00 (0.71 - 1.42) | 0.85 (0.59 - 1.21) |
| **Urban-Rural** |  |  |
| County Rural | 1.0 | 1.0 |
| County Suburban | 0.92 (0.80 - 1.06) | 0.97 (0.84 - 1.12) |
| County Urban | 0.66 (0.58 - 0.75)* | 0.88 (0.78 - 1.01) |
| **Education** |  |  |
| High School or Less | 1.0 | 1.0 |
| Some college | 0.94 (0.71 - 1.25) | 0.85 (0.64 - 1.14) |
| Associate Degree | 1.21 (0.91 - 1.60) | 0.95 (0.72 - 1.27) |
| Bachelor Degree | 0.87 (0.67 - 1.13) | 0.78 (0.59 - 1.02) |
| Graduate Degree | 0.64 (0.49 - 0.83) | 0.57 (0.44 - 0.76)* |
| **Influenza Vaccine History** |  |  |
| Never Had Influenza Vaccine | 1.0 | 1.0 |

| Term | Unadjusted Hazard Ratio (95% CI) | Adjusted Hazard Ratio (95% CI) |
| --- | --- | --- |
| Ever Had Influenza Vaccine | 0.78 (0.61 - 1.00) | 0.93 (0.72 - 1.20) |
| **Time Since Last Primary Care Visit** |  |  |
| Last Primary Care Visit < 1 Year Ago | 1.0 | 1.0 |
| Last Primary Care Visit 1-2 Years Ago | 0.94 (0.80 - 1.10) | 0.84 (0.71 - 0.98) * |
| Last Primary Care Visit 2-5 Years Ago | 0.84 (0.62 - 1.15) | 0.68 (0.49 - 0.94) * |
| Last Primary Care Visit >5 Years Ago | 1.32 (0.92 - 1.89) | 1.10 (0.77 - 1.59) |
| **Tobacco Use** |  |  |
| Not A Current Tobacco User | 1.0 | 1.0 |
| Current Tobacco User | 0.88 (0.70 - 1.11) | 0.73 (0.58 - 0.92) * |
| **Co-morbidities** |  |  |
| No Concurrent Medical Conditions | 1.0 | 1.0 |
| One Concurrent Medical Condition | 0.98 (0.87 - 1.10) | 1.01 (0.90 - 1.14) |
| Two Concurrent Medical Conditions | 0.91 (0.79 - 1.04) | 1.01 (0.88 - 1.17) |
| 3 or More Concurrent Medical Conditions | 0.99 (0.85 - 1.16) | 1.18 (0.99 - 1.40) |
| **Household Income** |  |  |
| Household Income <50k | 1.0 | 1.0 |
| Household income 50-100k | 1.27 (1.06 - 1.52) * | 1.22 (1.01 - 1.47) * |
| Household income >100k | 1.11 (0.93 - 1.32) | 1.14 (0.94 - 1.38) |
| Household income No answer | 0.96 (0.76 - 1.21) | 1.04 (0.82 - 1.33) |
| **In-person class** |  |  |
| No one in Household attends in person class | 1.0 | 1.0 |
| Someone in Household attends in person class | 1.64 (1.48 - 1.82) * | 1.28 (1.14 - 1.43) * |
| **Social Contact Occupation** |  |  |
| No one in Household works in a Social Contact Occupation | 1.0 | 1.0 |
| Someone in Household works in a Social Contact Occupation | 1.60 (1.45 - 1.77) * | 1.24 (1.12 - 1.38) * |
| **COVID-19 Vaccination** |  |  |

| Term | Unadjusted Hazard Ratio (95% CI) | Adjusted Hazard Ratio (95% CI) |
| --- | --- | --- |
| Has not yet had any COVID-19 Vaccine Doses | 1.0 | 1.0 |
| Had At Least One Vaccine Dose | 0.26 (0.23 - 0.30) * | 0.37 (0.32 - 0.42) * |
| **SOEM Risk Class** |  |  |
| SOEM - Class High Health Care Work | 1.0 | 1.0 |
| SOEM - Class High | 0.85 (0.74 - 0.98) * | 0.80 (0.69 - 0.93) * |
| SOEM Class Medium | 0.94 (0.78 - 1.13) * | 0.80 (0.66 - 0.97) * |
| SOEM - Class Low | 0.81 (0.69 - 0.94) * | 0.74 (0.63 - 0.87) * |
| SOEM Class No Data | 0.47 (0.41 - 0.54) * | 0.58 (0.49 - 0.68) * |

\* $p \leq 0.05$ ; Adjusted HR are adjusted for Sex, Age, Race-Ethnicity, Urban-Rural, Education, Influenza Vaccine History, Time Since Last Primary Care Visit, Tobacco Use, Co-morbidities, Household Income, In-person class, Social Contact Occupation, COVID-19 Vaccination and SOME Risk Class.

**Figure 1:**
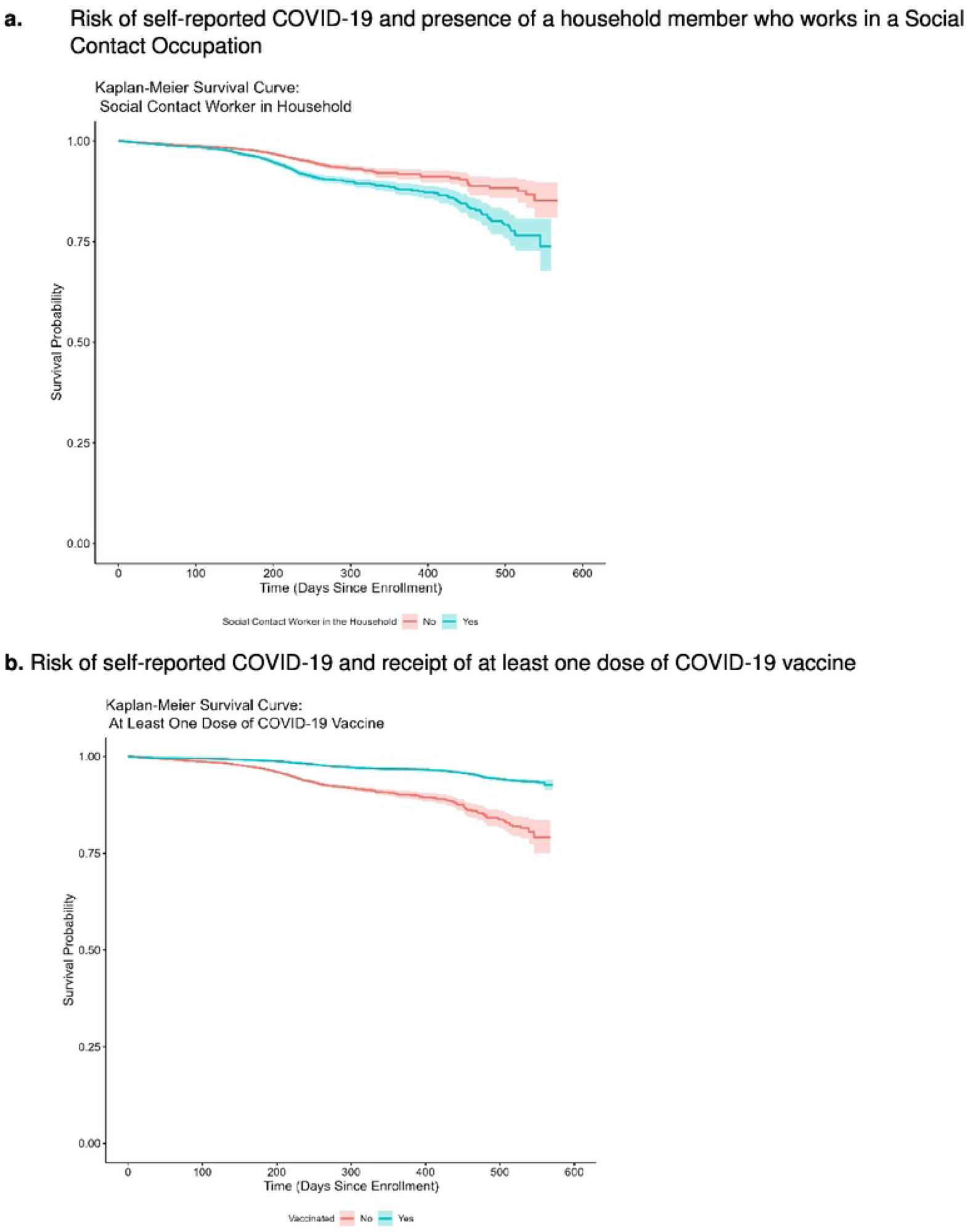
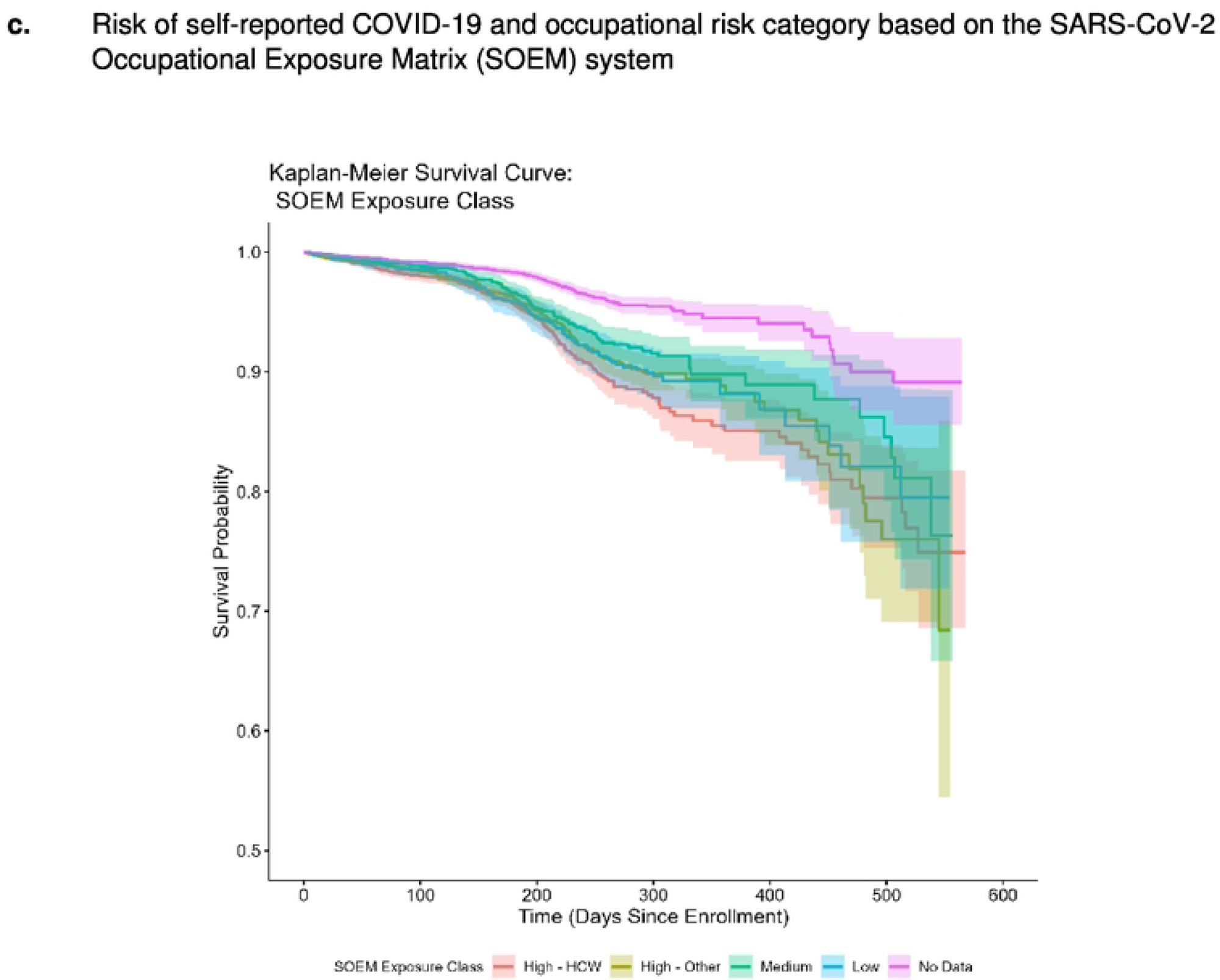
Kaplan Meyer survival analysis comparing cumulative risk of self-reported COVID-19 by specific risk factors.

## DISCUSSION

Our multicenter COVID-19 Community Research Partnership study provided a unique opportunity to examine the effect of demographic factors on risk of SARS CoV-2 infections in populations across the southern United States. After adjustment, we found that multiple social and economic factors were strongly associated with self-reported SARS CoV-2 infection during the pandemic. Risk of infection was significantly higher in young adult participants ages 18-34 years as compared to older groups, and the hazard ratios indicated that risk of infection as compared to the youngest participant group decreased further with each sequentially older age stratum, which is consistent with findings from other studies.^8,9^ This observation may be related to increasing concern about disease outcome in older age groups leading to greater adoption of preventative behaviors among older age cohorts, based on data that were well publicized in the media. In contrast to findings from other studies^10–12^, we found no association between race or ethnicity and risk of self-reported infection, a finding that may reflect the underrepresentation of minorities and the relative affluence of our study population. Participants with graduate level college education also demonstrated lower risk as compared to participants without college education, again consistent with other studies. ^13^ Potential explanations for this observation include a lower likelihood of work-related contact with the general public and a greater awareness of risk and effective methods of protection among more educated subjects. The observation of lower risk among those whose last encounter with a primary care provider 1-5 years ago as compared to those who saw their provider within the past year could be due to greater awareness of COVID-19 from the health care provider, or to health care visits related to COVID-19 illnesses.

Counterintuitively, smokers appeared to have a lower risk of infection as compared to non-smokers. This paradoxical relationship between smoking and risk of illness from COVID-19 has been observed in several other studies^14,15^, including some that speculate that nicotine could mitigate the effect of the cytokine storm in COVID-19 patients.^16^ However, these studies also point out that smokers are underrepresented in many COVID-19 population-based studies, as they appear to be in our study as well (5.4% of participants vs. the CDC estimated average of 12.4% for the southern United States in 2021)^17^, and the harmful effects of smoking are often cited as offsetting any possible benefit from smoking for COVID-19 related morbidity.

As expected, participants with higher occupational risk of exposure to the general public demonstrated higher risk of infection, including those living in a household with a person who attends classes in person or who encounter the general public in their workplace. When risk was compared by occupational group using the NIOSH SOEM risk categories, the most significant increase in risk was from working in a healthcare setting. These results may be used to inform identification of high-risk groups for future respiratory disease outbreaks, allowing targeted programs to promote protective measures and behaviors that reduce the risk of infection.

Our study is subject to several limitations. COVID-19 cases were ascertained based on self-reported data from daily surveys, so the infections could not be verified. Participants varied in the frequency with which they responded to the daily surveys, which limits our ability to determine the exact date of onset for the reported case of COVID-19. Generalizability is also limited due to the overrepresentation of lower risk socioeconomic groups in the study population and to recruitment that was limited to patients from healthcare networks and healthcare workers. As mentioned in the results section, this analysis was also limited to the 39% of participants who completed the supplemental questionnaires.

In contrast, the strengths of this study include a large sample size from a wide geographic area enrolled early in the pandemic and surveyed for a number of demographic, social, and economic characteristics. While our results illustrate the challenges inherent in understanding risk based on the complex interaction of age, race, and occupation and disentangling this from the risk due to local community infection rates, they also provide potential target groups for ongoing intervention. Under the assumption that those most at risk for first SARS-CoV-2 infection remain at higher risk for repeated infections even in the changing risk environment from vaccine uptake and shifting perceptions of the necessity of mitigations, this study may provide insight into ways to reduce ongoing disparities from the pandemic through risk stratification and targeted interventions.

## Data Availability

Data will be held in a public repository.

## Acknowledgements

The authors would like to thank the many dedicated CCRP participants who provided data for this study. All authors contributed to one or more areas of the methods (e.g. objectives, design, site selection and recruitment, form development, statistical approach) described in this article.

## Abbreviations and Acronyms

CCRP: COVID-19 Community Research Partnership
CDC: Centers for Disease Control and Prevention
HER: Electronic Health Records
IRB: Institutional Review Board

